# Using claims data to predict pre-operative BMI among bariatric surgery patients: development of the BMI Before Bariatric Surgery Scoring System (B3S3)

**DOI:** 10.1101/2023.10.04.23296517

**Authors:** Jenna Wong, Xiaojuan Li, David E. Arterburn, Dongdong Li, Elizabeth Messenger-Jones, Rui Wang, Sengwee Toh

## Abstract

**Background:** Lack of body mass index (BMI) measurements limits the utility of claims data for bariatric surgery research, but pre-operative BMI may be imputed due to existence of weight-related diagnosis codes and BMI-related reimbursements requirements. We used a machine learning pipeline to create a claims-based scoring system to predict pre-operative BMI, as documented in the electronic health record (EHR), among patients undergoing a new bariatric surgery.

**Methods:** Using the Optum Labs Data Warehouse, containing linked de-identified claims and EHR data for commercial or Medicare Advantage enrollees, we identified adults undergoing a new bariatric surgery between January 2011 and June 2018 with a BMI measurement in linked EHR data ≤30 days before the index surgery (n=3,226). We constructed predictors from claims data and applied a machine learning pipeline to create a scoring system for pre-operative BMI, the B3S3. We evaluated the B3S3 and a simple linear regression model (benchmark) in test patients whose index surgery occurred concurrent (2011-2017) or prospective (2018) to the training data.

**Results:** The machine learning pipeline yielded a final scoring system that included weight-related diagnosis codes, age, and number of days hospitalized and distinct drugs dispensed in the past 6 months. In concurrent test data, the B3S3 had excellent performance (R^2^ 0.862, 95% confidence interval [CI] 0.815-0.898) and calibration. The benchmark algorithm had good performance (R^2^ 0.750, 95% CI 0.686-0.799) and calibration but both aspects were inferior to the B3S3. Findings in prospective test data were similar.

**Conclusions:** The B3S3 is an accessible tool researchers can use with claims data to obtain granular and accurate predicted values of pre-operative BMI, which may enhance confounding control and investigation of effect modification by baseline obesity levels in bariatric surgery studies utilizing claims data.

## BACKGROUND

With a steady rise in the prevalence of severe obesity, defined as a body mass index (BMI) of 40 kg/m^2^ or greater, among US adults over the past two decades (age-adjusted prevalence of severe obesity increasing from 4.7% in 1999-2000 to 9.2% in 2017-2018),^1^ identifying effective and safe treatment options for severe obesity is an important public health priority. Compared to non-surgical treatments, bariatric surgery yields greater reductions in body weight, greater remission of chronic conditions like type 2 diabetes and metabolic syndrome, less medication use, and greater improvements in quality of life.^2,3^

Despite these benefits, more evidence on the comparative effectiveness and safety of different bariatric operations is needed, especially in the longer term. This need has been fueled by the rapid uptake of sleeve gastrectomy (SG) over the past decade, which has quickly displaced other previously common weight loss surgeries as the top bariatric operation in the United States, despite little supporting data.^4–6^ To address this gap, recent large-scale observational studies comparing post-surgical outcomes among different bariatric operations have largely utilized data from electronic health records (EHRs) or bariatric surgery registries.^5,7–10^ Administrative claims databases may be another viable and more cost-efficient data source for bariatric surgery research, offering the potential to identify larger and more representative patient populations and capture a more complete picture of patients’ medical encounters and post-surgical events across multiple health systems and over longer follow-up horizons.^6,11^

A concern of using claims databases for bariatric surgery research, however, relates to the ability to capture BMI data. This concern is warranted given that traditional claims databases lack clinical measurements,^11^ and weight-related diagnosis codes under the International Classification of Diseases (ICD) system are severely underutilized in most patient populations.^12–14^ However, bariatric surgery patients, particularly in the pre-operative period, may be an exception because most US health insurers require documentation of a BMI ≥40 kg/m^2^ or ≥35 kg/m^2^ with an obesity-related comorbidity for patients to receive prior approval and coverage of their operations.^15^ This requirement incentivizes documentation of weight-related ICD codes for virtually all (>98%) bariatric surgery patients,^16^ creating a unique opportunity for claims data to contain valuable information on pre-operative BMI. An important potential confounder of bariatric surgery comparative effectiveness studies, pre-operative BMI has been found to both vary across patients undergoing different bariatric operations^5,7,16,17^ and impact the risk of numerous safety and effectiveness outcomes, including post-operative complications,^17–20^ mortality,^20–22^ weight loss,^17,23,24^ and type 2 diabetes remission.^25,26^

A previous study^16^ found that weight codes in claims data during the pre-operative period had excellent concordance with pre-operative BMI in the EHR for identifying bariatric patients with a BMI of ≥35 kg/m^2^ at baseline. However, as more granular measures of BMI are desired, such high concordance is increasingly challenging to attain due to differences in the precision of weight-related ICD codes (that can only denote BMI *ranges*, e.g., 45.0-49.9 kg/m^2^, or nonspecific weight categories, e.g., obesity) versus numeric BMI values in the EHR (e.g., 47.2 kg/m^2^), as well as potential inaccuracies in documentation of pre-operative weight codes due to reasons such as miscoding (e.g., data entry errors), differential coding (e.g., unequal documentation of BMI-specific versus nonspecific weight codes across operations), or weight loss before the operation (e.g., as part of the patient’s treatment plan).^16^

Rather than infer pre-operative BMI from the descriptions of weight-related ICD codes, two enhancements may allow for more precise and accurate prediction of pre-operative BMI from claims data: 1) use of additional predictors (i.e., features) from claims data, besides weight-related diagnosis codes, and 2) use of flexible, data-driven modeling techniques to predict pre-operative BMI. However, for such prediction models to be of practical use in bariatric surgery research, they must also be easy to disseminate and for investigators to interpret and implement in claims databases. In this study, we applied a data-driven machine learning pipeline that considered a broad range of features from claims data to create and internally validate an accessible scoring system to predict pre-operative BMI, as documented in the EHR, among bariatric surgery patients.

## METHODS

### Data Source

This study used data from the Optum Labs Data Warehouse (OLDW), containing linked de-identified administrative claims and EHR data for commercially insured and Medicare Advantage enrollees. The database contains longitudinal health information on enrollees and patients, representing a diverse mixture of ages, races/ethnicities, and geographical regions across the United States.^27^ The claims component includes physician, pharmacy, and facility claims submitted for covered enrollees, while the EHR component includes clinical diagnoses, procedures, prescriptions, clinical notes, laboratory results, and vital signs (including BMI) documented for enrollees as part of routine clinical practice.

This study was approved by the Harvard Pilgrim Health Care Institutional Review Board with a waiver of individual patient consent.

### Study Population

Following the inclusion and exclusion criteria from a previous study,^16^ we used the claims portion of the OLDW to identify a retrospective cohort of patients aged ≥18 years who newly underwent SG, Roux-en-Y gastric bypass (RYGB), or adjusted gastric banding (AGB) for weight loss between January 1, 2011 and June 30, 2018. The index bariatric operation could occur in an inpatient or ambulatory care setting, and eligible patients required continuous medical and pharmacy coverage during the 6-month period before their index surgery (see eFigure 1 in Supplementary File 1 for more details). From this eligible cohort, we identified the subset of patients with a BMI measurement recorded in their linked EHR data on or within 30 days prior to the day of the index surgery, where we used this subset of patients to derive and validate the scoring system for pre-operative BMI.

**Figure 1.**
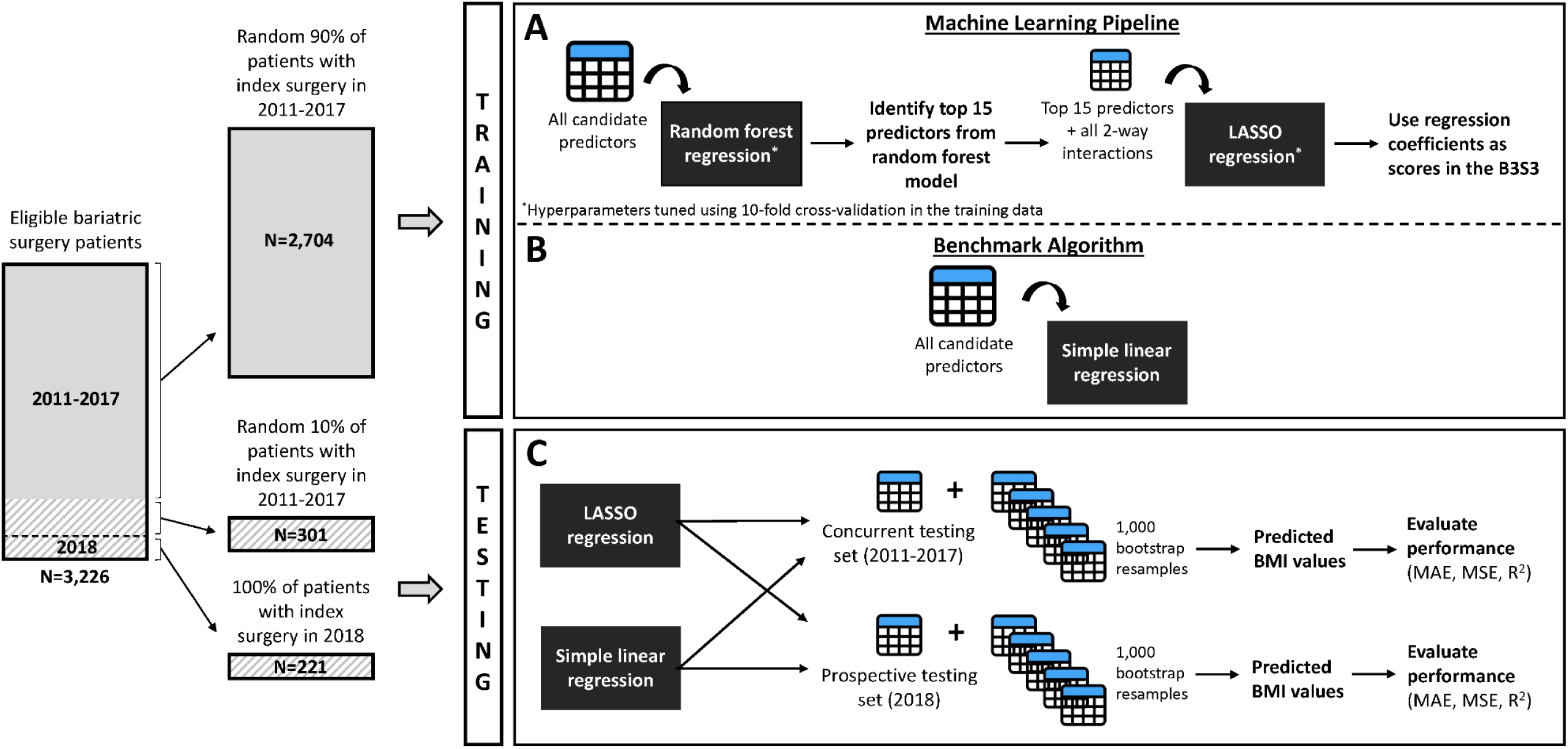
Derivation and validation of the B3S3 and benchmark algorithm. Abbreviations: B3S3 = BMI Before Bariatric Surgery Scoring System; LASSO = least absolute shrinkage and selection operator; MAE = mean absolute error; MSE = mean squared error The training data (solid grey box) were used to derive two prediction models for pre-operative BMI: 1) a LASSO regression model created through a machine learning pipeline, where a random forest regression model was first used to identify the 15 most important features among all candidate predictors, and then the top 15 features and all 2-way interactions between them were used as inputs into a LASSO regression model (Panel A), and 2) a simple linear regression model utilizing all candidate predictors as inputs without any variable selection or interaction terms (Panel B). The performance of the LASSO model and the simple linear regression model, fit on the full training data, was evaluated in two hold-out sets of the study data not used during training (the concurrent and prospective testing set; hatched grey boxes). The final fitted models were also evaluated in 1,000 bootstrap resamples of the 2 testing sets to calculate empirical 95% confidence intervals for all performance metrics (Panel C). The regression coefficients from the fitted LASSO model represent the scores in the B3S3.

### Model Outcome

We used the BMI values recorded in the EHR (“EHR-based”) on or within 30 days prior to the date of the index surgery as the reference standard measure of pre-operative BMI and the model outcome. All EHR-based BMI measurements were recorded as a continuous value, which we expressed to the nearest tenth of a unit (e.g., 47.2 kg/m^2^). If multiple BMI measurements were recorded for the patient within the 30-day pre-operative period, we used the measurement closest to the index surgery.

### Candidate Predictors

From the claims portion of the OLDW, we used data in the 6 months prior to the index surgery to create various domains of candidate features for pre-operative BMI (“claims-based”). The first domain included indicators for 14 weight code categories, 12 of which captured ICD codes for specific BMI categories and 2 of which captured ICD codes for non-specific weight categories (see eTable 1 in Supplementary File 2 for category descriptions and code mappings). These indicators were used to capture all weight-related codes documented for a given patient in the prior 6 months. Other domains of claims-based features included patient demographics (age, sex, race/ethnicity, geographical region within the US, and insurance type), comorbidities (33 indicators of conditions in the Charlson/Elixhauser combined comorbidity score^28,29^ and conditions strongly associated with obesity, such as gastroesophageal reflux disease, sleep apnea, dyslipidemia, and acquired hypothyroidism), hospitalizations in the past 6 months (number of hospitalizations and total days hospitalized), dispensed medications in the past 6 months (total number of distinct generic drugs, indicator for medications approved for management of overweight and obesity, and indicators for 6 medication classes with weight gain as a known side effect), and the bariatric surgery type (SG, RYBG, or AGB).

### Machine Learning Pipeline

Using a randomly selected 90% of study patients whose index operation occurred in 2011-2017 (training set), we applied a machine learning pipeline designed to yield an accurate, yet also accessible tool to predict pre-operative BMI from claims data (Figure 1, Panel A).

The pipeline involved first training a random forest regression model using all candidate features as inputs to predict pre-operative BMI. A nonparametric ensemble machine learning technique, the random forest methodology^30^ can naturally accommodate interactions and nonlinear associations in the data without any manual model specification (making it more robust to model misspecification) and is more resilient to problems of high variability and overfitting that often plague the use of single decision trees.^31,32^ Despite these advantages and the often superior performance of the random forest algorithm over other machine learning techniques,^32^ this algorithm is also more complex to interpret and cumbersome to disseminate, creating practical barriers for such models to be shared and implemented by researchers.

Thus, in the second part of the pipeline, we attempted to transfer the insights learned by the random forest model to a more familiar and accessible type of prediction algorithm – a linear regression model. We ranked the features in the random forest model in decreasing order of variable importance (measured by the total decrease in node impurities, or residual sum of squares, from splitting on a given variable, averaged over all trees in the random forest^33^) and considered for inclusion in a linear regression model the top 15 predictors, along with all 2-way interactions between them to capture any 2-way interactions that may have been modeled in the random forest model. We selected a threshold of the top 15 predictors in light of the size of the training set to avoid overparameterization of the linear regression model after including all 2-way interactions. We offered these variables to a least absolute shrinkage and selection operator (LASSO) regression model, which applied shrinkage to the model coefficients (to reduce model variability) and performed automated variable selection (by shrinking some coefficients right to zero). The fitted LASSO model represented the final prediction model, and its coefficients represented the points in our scoring system, which we refer to as the *BMI Before Bariatric Surgery Scoring System* (B3S3). For both the random forest and LASSO algorithms, we tuned their hyperparameter values using a grid search procedure that iteratively assessed the algorithms’ cross-validated performance over a range of plausible values (see eTable 2 in Supplementary File 2 for details and the tuned values). Although it is typically recommended to first scale the value of continuous variables in a LASSO model, we found that in our case, scaling had no impact on the LASSO model’s performance and therefore used the original values to facilitate the use and interpretation of the scoring system.

### Benchmark Algorithm

We compared the performance of the LASSO model (i.e., the B3S3), produced through our machine learning pipeline, to a simple linear regression model with all candidate predictors as inputs and no variable selection or interaction terms (Figure 1, Panel B). Thus, the simple linear regression model represented a benchmark algorithm that was offered the same set of features as the machine learning pipeline but whose model specification was not determined through a flexible, machine-guided process.

### Performance Evaluation

We evaluated (internally validated) the performance of the B3S3 and the benchmark algorithm by applying these models in 2 hold-out sets of the study data that were not used for training (Figure 1, Panel C). The first set included the remaining 10% of study patients whose index operation occurred in 2011-2017 (concurrent testing set). The second set included all study patients whose index operation occurred in 2018 (prospective testing set).

We assessed overall model performance using the following metrics: 1) mean absolute error (MAE), where the error was the absolute difference between the predicted and observed pre-operative BMI value, 2) mean squared error (MSE), where the absolute error was squared to amplify observations with larger errors and 3) R^2^, which quantified the proportion of variance in observed pre-operative BMI values explained by the model, where R^2^ values closer to 1 indicated better overall performance.

We assessed model calibration (i.e., accuracy of the model predictions) numerically and visually. We calculated the ratio of the mean predicted to observed pre-operative BMI, first overall (“weak calibration”) and then within 3 strata of predicted BMI: <40.0, 40.0-<50.0, ≥50.0 kg/m^2^ (“moderate calibration”)^34^, where ratios closer to 1 indicated better calibration. We also constructed calibration plots of observed versus predicted BMI values, where well-calibrated observations fell along the diagonal (indicating perfect agreement between the predicted and observed BMI values). To maintain the de-identification nature of the database, we coarsened the calibration plot into 5-unit BMI categories and masked areas of the plot with <11 patients.

To measure variability in model performance due to the sampling procedure used to create the testing sets, we applied the final fitted models to 1,000 bootstrap resamples of the testing sets and reported the values at the 2.5^th^ and 97.5^th^ percentiles of the resulting distribution.

All analyses were performed in the R environment for statistical computing, version 4.0.2. We used the glm (v4.0.2), glmnet (v4.0.2), and randomForest (v4.6.14) packages to fit the linear, LASSO, and random forest regression models, respectively.

## RESULTS

A total of 3,226 bariatric surgery patients met the study inclusion criteria. Most patients were female (76.3%), middle-aged (median 48 years old), and had commercial health insurance (77.6%). The race/ethnicity of most patients was white (73.0%), followed by black (15.7%) and Hispanic (7.0%), and most patients were from the Midwest (54.0%) or South (33.6%).

Hypertension, gastroesophageal reflux disease, sleep apnea, and dyslipidemia were the most common baseline comorbidities, with each affecting more than 50% of study patients (see eTable 3 in Supplementary File 2 for the prevalence of all 33 comorbidities considered for the scoring system). By far, the most common bariatric operation was SG (58.2%), followed by RYGB (35.2%) and AGB (6.6%). SG was performed more frequently in 2018 than in 2011-2017 (Table 1).

**Table 1.**
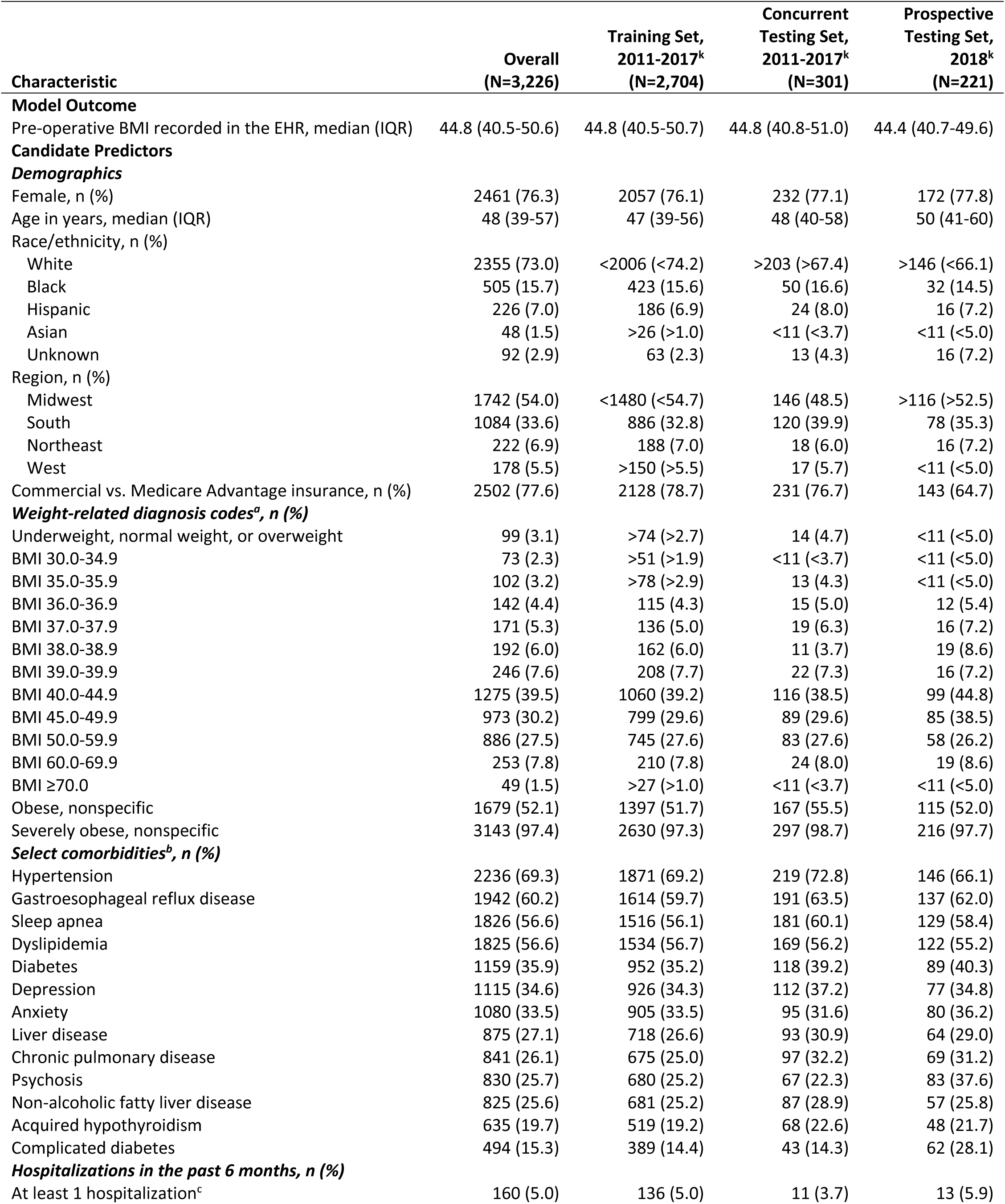
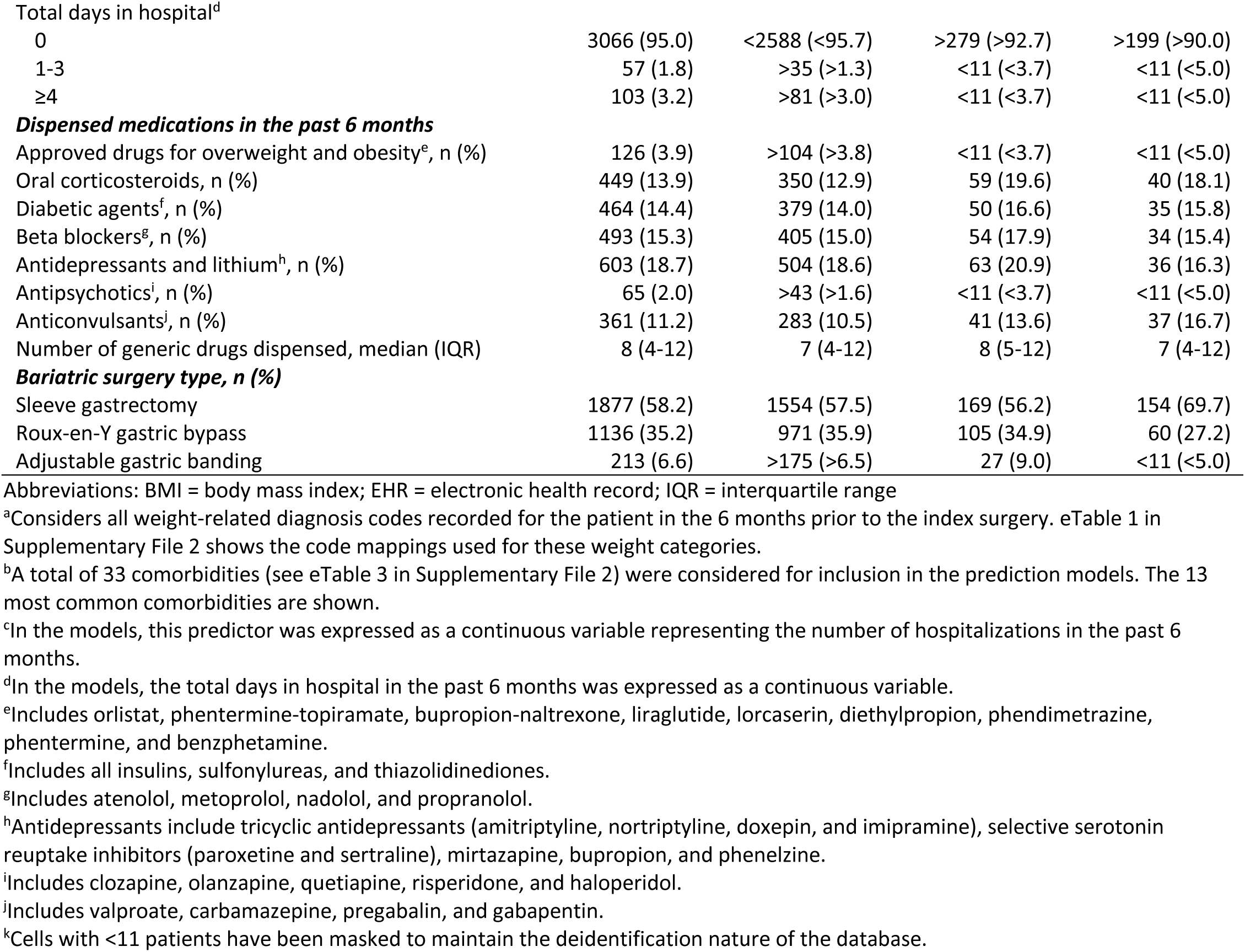
Characteristics of the study cohort.

| Characteristic | Overall<br>(N=3,226) | Training Set,<br>2011-2017<sup>k</sup><br>(N=2,704) | Concurrent<br>Testing Set,<br>2011-2017<sup>k</sup><br>(N=301) | Prospective<br>Testing Set,<br>2018<sup>k</sup><br>(N=221) |
| --- | --- | --- | --- | --- |
| **Model Outcome** |  |  |  |  |
| Pre-operative BMI recorded in the EHR, median (IQR) | 44.8 (40.5-50.6) | 44.8 (40.5-50.7) | 44.8 (40.8-51.0) | 44.4 (40.7-49.6) |
| **Candidate Predictors** |  |  |  |  |
| **Demographics** |  |  |  |  |
| Female, n (%) | 2461 (76.3) | 2057 (76.1) | 232 (77.1) | 172 (77.8) |
| Age in years, median (IQR) | 48 (39-57) | 47 (39-56) | 48 (40-58) | 50 (41-60) |
| Race/ethnicity, n (%) |  |  |  |  |
| White | 2355 (73.0) | <2006 (<74.2) | >203 (>67.4) | >146 (<66.1) |
| Black | 505 (15.7) | 423 (15.6) | 50 (16.6) | 32 (14.5) |
| Hispanic | 226 (7.0) | 186 (6.9) | 24 (8.0) | 16 (7.2) |
| Asian | 48 (1.5) | >26 (>1.0) | <11 (<3.7) | <11 (<5.0) |
| Unknown | 92 (2.9) | 63 (2.3) | 13 (4.3) | 16 (7.2) |
| Region, n (%) |  |  |  |  |
| Midwest | 1742 (54.0) | <1480 (<54.7) | 146 (48.5) | >116 (>52.5) |
| South | 1084 (33.6) | 886 (32.8) | 120 (39.9) | 78 (35.3) |
| Northeast | 222 (6.9) | 188 (7.0) | 18 (6.0) | 16 (7.2) |
| West | 178 (5.5) | >150 (>5.5) | 17 (5.7) | <11 (<5.0) |
| Commercial vs. Medicare Advantage insurance, n (%) | 2502 (77.6) | 2128 (78.7) | 231 (76.7) | 143 (64.7) |
| **Weight-related diagnosis codes<sup>a</sup>, n (%)** |  |  |  |  |
| Underweight, normal weight, or overweight | 99 (3.1) | >74 (>2.7) | 14 (4.7) | <11 (<5.0) |
| BMI 30.0-34.9 | 73 (2.3) | >51 (>1.9) | <11 (<3.7) | <11 (<5.0) |
| BMI 35.0-35.9 | 102 (3.2) | >78 (>2.9) | 13 (4.3) | <11 (<5.0) |
| BMI 36.0-36.9 | 142 (4.4) | 115 (4.3) | 15 (5.0) | 12 (5.4) |
| BMI 37.0-37.9 | 171 (5.3) | 136 (5.0) | 19 (6.3) | 16 (7.2) |
| BMI 38.0-38.9 | 192 (6.0) | 162 (6.0) | 11 (3.7) | 19 (8.6) |
| BMI 39.0-39.9 | 246 (7.6) | 208 (7.7) | 22 (7.3) | 16 (7.2) |
| BMI 40.0-44.9 | 1275 (39.5) | 1060 (39.2) | 116 (38.5) | 99 (44.8) |
| BMI 45.0-49.9 | 973 (30.2) | 799 (29.6) | 89 (29.6) | 85 (38.5) |
| BMI 50.0-59.9 | 886 (27.5) | 745 (27.6) | 83 (27.6) | 58 (26.2) |
| BMI 60.0-69.9 | 253 (7.8) | 210 (7.8) | 24 (8.0) | 19 (8.6) |
| BMI ≥70.0 | 49 (1.5) | >27 (>1.0) | <11 (<3.7) | <11 (<5.0) |
| Obese, nonspecific | 1679 (52.1) | 1397 (51.7) | 167 (55.5) | 115 (52.0) |
| Severely obese, nonspecific | 3143 (97.4) | 2630 (97.3) | 297 (98.7) | 216 (97.7) |
| **Select comorbidities<sup>b</sup>, n (%)** |  |  |  |  |
| Hypertension | 2236 (69.3) | 1871 (69.2) | 219 (72.8) | 146 (66.1) |
| Gastroesophageal reflux disease | 1942 (60.2) | 1614 (59.7) | 191 (63.5) | 137 (62.0) |
| Sleep apnea | 1826 (56.6) | 1516 (56.1) | 181 (60.1) | 129 (58.4) |
| Dyslipidemia | 1825 (56.6) | 1534 (56.7) | 169 (56.2) | 122 (55.2) |
| Diabetes | 1159 (35.9) | 952 (35.2) | 118 (39.2) | 89 (40.3) |
| Depression | 1115 (34.6) | 926 (34.3) | 112 (37.2) | 77 (34.8) |
| Anxiety | 1080 (33.5) | 905 (33.5) | 95 (31.6) | 80 (36.2) |
| Liver disease | 875 (27.1) | 718 (26.6) | 93 (30.9) | 64 (29.0) |
| Chronic pulmonary disease | 841 (26.1) | 675 (25.0) | 97 (32.2) | 69 (31.2) |
| Psychosis | 830 (25.7) | 680 (25.2) | 67 (22.3) | 83 (37.6) |
| Non-alcoholic fatty liver disease | 825 (25.6) | 681 (25.2) | 87 (28.9) | 57 (25.8) |
| Acquired hypothyroidism | 635 (19.7) | 519 (19.2) | 68 (22.6) | 48 (21.7) |
| Complicated diabetes | 494 (15.3) | 389 (14.4) | 43 (14.3) | 62 (28.1) |
| **Hospitalizations in the past 6 months, n (%)** |  |  |  |  |
| At least 1 hospitalization<sup>c</sup> | 160 (5.0) | 136 (5.0) | 11 (3.7) | 13 (5.9) |
| Total days in hospital<sup>d</sup> |  |  |  |  |
| 0 | 3066 (95.0) | <2588 (<95.7) | >279 (>92.7) | >199 (>90.0) |
| 1-3 | 57 (1.8) | >35 (>1.3) | <11 (<3.7) | <11 (<5.0) |
| ≥4 | 103 (3.2) | >81 (>3.0) | <11 (<3.7) | <11 (<5.0) |
| **Dispensed medications in the past 6 months** |  |  |  |  |
| Approved drugs for overweight and obesity<sup>e</sup>, n (%) | 126 (3.9) | >104 (>3.8) | <11 (<3.7) | <11 (<5.0) |
| Oral corticosteroids, n (%) | 449 (13.9) | 350 (12.9) | 59 (19.6) | 40 (18.1) |
| Diabetic agents<sup>f</sup>, n (%) | 464 (14.4) | 379 (14.0) | 50 (16.6) | 35 (15.8) |
| Beta blockers<sup>g</sup>, n (%) | 493 (15.3) | 405 (15.0) | 54 (17.9) | 34 (15.4) |
| Antidepressants and lithium<sup>h</sup>, n (%) | 603 (18.7) | 504 (18.6) | 63 (20.9) | 36 (16.3) |
| Antipsychotics<sup>i</sup>, n (%) | 65 (2.0) | >43 (>1.6) | <11 (<3.7) | <11 (<5.0) |
| Anticonvulsants<sup>j</sup>, n (%) | 361 (11.2) | 283 (10.5) | 41 (13.6) | 37 (16.7) |
| Number of generic drugs dispensed, median (IQR) | 8 (4-12) | 7 (4-12) | 8 (5-12) | 7 (4-12) |
| **Bariatric surgery type, n (%)** |  |  |  |  |
| Sleeve gastrectomy | 1877 (58.2) | 1554 (57.5) | 169 (56.2) | 154 (69.7) |
| Roux-en-Y gastric bypass | 1136 (35.2) | 971 (35.9) | 105 (34.9) | 60 (27.2) |
| Adjustable gastric banding | 213 (6.6) | >175 (>6.5) | 27 (9.0) | <11 (<5.0) |

Abbreviations: BMI = body mass index; EHR = electronic health record; IQR = interquartile range
<sup>a</sup>Considers all weight-related diagnosis codes recorded for the patient in the 6 months prior to the index surgery. eTable 1 in Supplementary File 2 shows the code mappings used for these weight categories.
<sup>b</sup>A total of 33 comorbidities (see eTable 3 in Supplementary File 2) were considered for inclusion in the prediction models. The 13 most common comorbidities are shown.
<sup>c</sup>In the models, this predictor was expressed as a continuous variable representing the number of hospitalizations in the past 6 months.
<sup>d</sup>In the models, the total days in hospital in the past 6 months was expressed as a continuous variable.
<sup>e</sup>Includes orlistat, phentermine-topiramate, bupropion-naltrexone, liraglutide, lorcaserin, diethylpropion, phendimetrazine, phentermine, and benzphetamine.
<sup>f</sup>Includes all insulins, sulfonylureas, and thiazolidinediones.
<sup>g</sup>Includes atenolol, metoprolol, nadolol, and propranolol.
<sup>h</sup>Antidepressants include tricyclic antidepressants (amitriptyline, nortriptyline, doxepin, and imipramine), selective serotonin reuptake inhibitors (paroxetine and sertraline), mirtazapine, bupropion, and phenelzine.
<sup>i</sup>Includes clozapine, olanzapine, quetiapine, risperidone, and haloperidol.
<sup>j</sup>Includes valproate, carbamazepine, pregabalin, and gabapentin.
<sup>k</sup>Cells with <11 patients have been masked to maintain the deidentification nature of the database.

The median (interquartile range) pre-operative BMI in the EHR was 44.8 (40.5-50.6) kg/m^2^, which was documented on the day of the index operation for over 50% of patients. In linked claims data during the 6-month period before the index operation, 98.3% of patients had a weight-related ICD code documented, and 96.0% had a BMI-specific code documented. Non-specific weight codes for severe obesity were more commonly documented than non-specific codes for obesity (97.4% versus 52.1%), while the top documented BMI-specific weight codes were for the ranges 40.0-44.9 kg/m^2^ (39.5%) and 45.0-49.9 kg/m^2^ (30.2%), which appeared more frequently for surgeries in 2018 than in 2011-2017 (Table 1).

### Machine Learning Pipeline

The fitted random forest model identified the following 15 variables as most important: 12 indicators for weight-related code categories and 3 continuous variables for patient age (ranked 7^th^), number of distinct generic drugs dispensed in the past 6 months (ranked 12^th^), and number of days hospitalized in the past 6 months (ranked 15^th^). In ascending rank order, the top 3 predictors were indicators for the following weight code categories: “BMI 60.0-69.9 kg/m^2^”, “BMI 50.0-59.9 kg/m^2^”, and “Severely obese, non-specific”. Absent from the top 15 predictors were the weight code categories “Underweight, normal weight, or overweight” and “Obese, nonspecific”, as well as features related to comorbidities or previously dispensed medications.

When the top 15 predictors and all their 2-way interactions were offered to a LASSO regression model (a total of 120 terms), the fitted model retained 68 terms, of which 57 were 2-way interactions that largely included interactions between weight code categories, effectively adjusting the predicted BMI in cases where diagnosis codes for multiple weight categories (e.g., conflicting categories or BMI-specific and non-specific weight categories) were documented for the patient in the previous 6 months. The coefficients of the LASSO model ranged in magnitude from −26.362 to 29.480 (Table 2) and were used as the scores in the B3S3. Supplementary File 3 contains an Excel spreadsheet showing step-by-step instructions on how to use the B3S3, along with an interactive example.

**Table 2.** Coefficients of the LASSO regression model.

| **Variable** | **Regression coefficient<sup>a</sup>** |
| --- | --- |
| Intercept | 28.141 |
| ***Individual predictors*** |  |
| Code for BMI 36.0-36.9 | -4.289 |
| Code for BMI 38.0-38.9 | -3.113 |
| Code for BMI 40.0-44.9 | -0.377 |
| Code for BMI 45.0-49.9 | 3.329 |
| Code for BMI 50.0-59.9 | 10.371 |
| Code for BMI 60.0-69.9 | 17.795 |
| Code for BMI 70.0+ | 29.480 |
| Code for severely obese, nonspecific | 15.750 |
| Age in years | -0.025 |
| Number of distinct generic drugs dispensed in the past 6 months | -0.010 |
| Total days hospitalized in the past 6 months | -0.029 |
| ***2-way interactions between weight-related codes*** |  |
| Code for BMI 30.0-34.9 x Code for BMI 35.0-35.9 | 2.908 |
| Code for BMI 30.0-34.9 x Code for BMI 36.0-36.9 | 1.814 |
| Code for BMI 30.0-34.9 x Code for BMI 37.0-37.9 | 3.074 |
| Code for BMI 30.0-34.9 x Code for BMI 38.0-38.9 | 2.324 |
| Code for BMI 30.0-34.9 x Code for BMI 39.0-39.9 | 2.001 |
| Code for BMI 30.0-34.9 x Code for BMI 40.0-44.9 | 2.471 |
| Code for BMI 30.0-34.9 x Code for BMI 45.0-49.9 | 3.536 |
| Code for BMI 30.0-34.9 x Code for BMI 50.0-59.9 | 2.935 |
| Code for BMI 30.0-34.9 x Code for BMI 60.0-69.9 | -19.947 |
| Code for BMI 30.0-34.9 x Code for severely obese, nonspecific | -6.458 |
| Code for BMI 35.0-35.9 x Code for BMI 36.0-36.9 | 1.719 |
| Code for BMI 35.0-35.9 x Code for BMI 37.0-37.9 | 0.941 |
| Code for BMI 35.0-35.9 x Code for BMI 38.0-38.9 | 1.131 |
| Code for BMI 35.0-35.9 x Code for BMI 39.0-39.9 | 1.205 |
| Code for BMI 35.0-35.9 x Code for BMI 40.0-44.9 | 1.171 |
| Code for BMI 35.0-35.9 x Code for BMI 45.0-49.9 | 1.848 |
| Code for BMI 35.0-35.9 x Code for severely obese, nonspecific | -4.299 |
| Code for BMI 36.0-36.9 x Code for BMI 37.0-37.9 | 1.424 |
| Code for BMI 36.0-36.9 x Code for BMI 38.0-38.9 | 1.637 |
| Code for BMI 36.0-36.9 x Code for BMI 39.0-39.9 | 0.201 |
| Code for BMI 36.0-36.9 x Code for BMI 50.0-59.9 | -11.079 |
| Code for BMI 37.0-37.9 x Code for BMI 38.0-38.9 | 1.135 |
| Code for BMI 37.0-37.9 x Code for BMI 39.0-39.9 | 0.520 |
| Code for BMI 37.0-37.9 x Code for BMI 40.0-44.9 | 0.728 |
| Code for BMI 37.0-37.9 x Code for BMI 45.0-49.9 | -1.560 |
| Code for BMI 37.0-37.9 x Code for severely obese, nonspecific | -2.889 |
| Code for BMI 38.0-38.9 x Code for BMI 39.0-39.9 | 1.911 |
| Code for BMI 38.0-38.9 x Code for BMI 40.0-44.9 | 0.098 |
| Code for BMI 38.0-38.9 x Code for BMI 45.0-49.9 | -4.010 |
| Code for BMI 38.0-38.9 x Code for BMI 50.0-59.9 | -2.485 |
| Code for BMI 39.0-39.9 x Code for BMI 45.0-49.9 | 0.063 |
| Code for BMI 39.0-39.9 x Code for severely obese, nonspecific | -2.259 |
| Code for BMI 40.0-44.9 x Code for BMI 45.0-49.9 | -1.835 |
| Code for BMI 40.0-44.9 x Code for BMI 50.0-59.9 | -2.348 |
| Code for BMI 40.0-44.9 x Code for BMI 70.0+ | -26.362 |
| Code for BMI 40.0-44.9 x Code for severely obese, nonspecific | -0.002 |
| Code for BMI 45.0-49.9 x Code for BMI 50.0-59.9 | -8.077 |
| Code for BMI 45.0-49.9 x Code for BMI 60.0-69.9 | -14.917 |
| Code for BMI 45.0-49.9 x Code for BMI 70.0+ | -3.649 |
| Code for BMI 45.0-49.9 x Code for severely obese, nonspecific | 0.094 |
| Code for BMI 50.0-59.9 x Code for BMI 60.0-69.9 | -14.423 |
| Code for BMI 50.0-59.9 x Code for BMI 70.0+ | -15.892 |
| Code for BMI 60.0-69.9 x Code for BMI 70.0+ | -24.218 |
| Code for BMI 60.0-69.9 x Code for severely obese, nonspecific | 0.170 |
| Code for BMI 70.0+ x Code for severely obese, nonspecific | 0.402 |
| ***2-way interactions between weight-related codes and age*** |  |
| Code for BMI 45.0-49.9 x Age in years | 0.014 |
| Code for BMI 50.0-59.9 x Age in years | 0.008 |
| Code for BMI 60.0-69.9 x Age in years | 0.035 |
| ***2-way interactions between weight-related codes and number of distinct generic drugs dispensed in the past 6 months*** |  |
| Code for BMI 37.0-37.9 x Number of generic drugs | -0.008 |
| Code for BMI 45.0-49.9 x Number of generic drugs | 0.005 |
| ***2-way interactions between weight-related codes and total days hospitalized in the past 6 months*** |  |
| Code for BMI 30.0-34.9 x Total days hospitalized | 0.007 |
| Code for BMI 39.0-39.9 x Total days hospitalized | 0.079 |
| Code for BMI 50.0-59.9 x Total days hospitalized | 0.197 |
| Code for BMI 60.0-69.9 x Total days hospitalized | 0.062 |
| Code for BMI 70.0+ x Total days hospitalized | -0.577 |
| Code for severely obese, nonspecific x Total days hospitalized | 0.005 |
| ***2-way interactions between predictors besides weight-related codes*** |  |
| Age in years x Total days hospitalized | -0.001 |

Abbreviations: BMI = body mass index; LASSO = least absolute shrinkage and selection operator
<sup>a</sup>Represents the scores in the BMI Before Bariatric Surgery Scoring System (B3S3). Supplementary File 3 contains a spreadsheet with step-by-step instructions on how to use the B3S3, along with an interactive example.

### Overall Performance of the B3S3 and Benchmark Algorithm

In the concurrent testing set, the overall performance of the B3S3 was excellent, with a MAE of 2.309 (95% CI 2.045-2.575) (indicating that the prediction for a given patient was, on average, 2.309 BMI units different from the patient’s pre-operative BMI documented in the EHR), MSE of 10.682 (95% CI 7.854-13.584), and R^2^ of 0.862 (95% CI 0.815-0.898). In comparison, the overall performance of the simple linear regression model (benchmark algorithm) was good, but inferior to the B3S3, with a higher MAE of 3.266 (95% CI 2.933-3.610), higher MSE of 19.306 (95% CI 15.274-23.983) and lower R^2^ of 0.750 (95% CI 0.686-0.799), where all 95% CIs were non-overlapping with the B3S3. Findings were similar in the prospective testing set (Table 3).

**Table 3.**
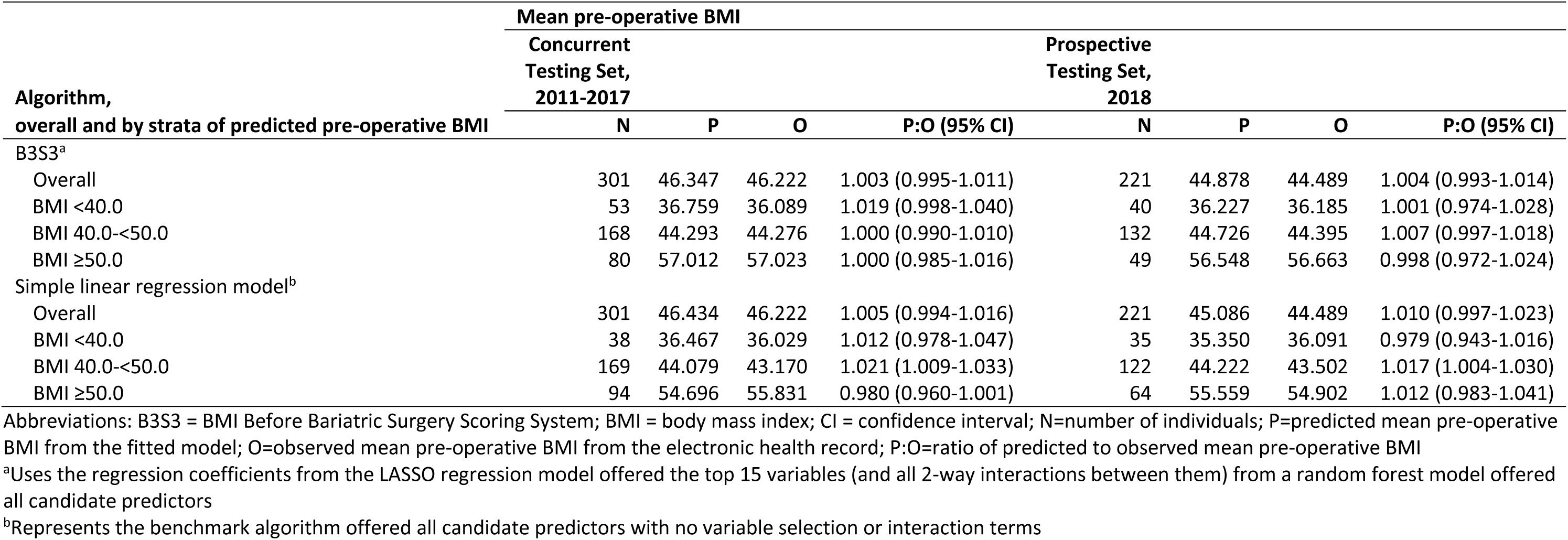
Performance of the B3S3 and the simple linear regression model.

### Calibration of the B3S3 and Benchmark Algorithm

In the concurrent testing set, the overall mean predicted pre-operative BMI from the B3S3 and simple linear regression model differed from the overall mean observed pre-operative BMI by only 0.3% (95% CI −0.5%-1.1%) and 0.5% (95% CI −0.6%-1.6%), respectively. When patients were grouped into more homogeneous strata based on their predicted BMI (<40.0, 40.0-<50.0, ≥50.0 kg/m^2^), the calibration of the B3S3 was still excellent, with the mean predicted and observed BMI differing by <2% in the lowest stratum and <0.05% in the upper 2 strata, where the 95% CI around the predicted to observed ratio for all strata included 1, indicating no significant differences (Table 4). For the simple linear regression model, however, the mean predicted and observed values for the upper 2 BMI strata differed by >2%, with the 95% CI around the predicted to observed ratio for the middle (and most common) BMI strata being above 1, indicating that the model tended to slightly overestimate pre-operative BMI for predictions in this range. Findings were similar in the prospective testing set (Table 4). The calibration plot further illustrated the slightly better accuracy of predictions from the B3S3, where patients in both testing sets were more tightly and evenly scattered along the diagonal for the B3S3 (Figure 2, top row) than the simple linear regression model (Figure 2, bottom row).

**Figure 2.**
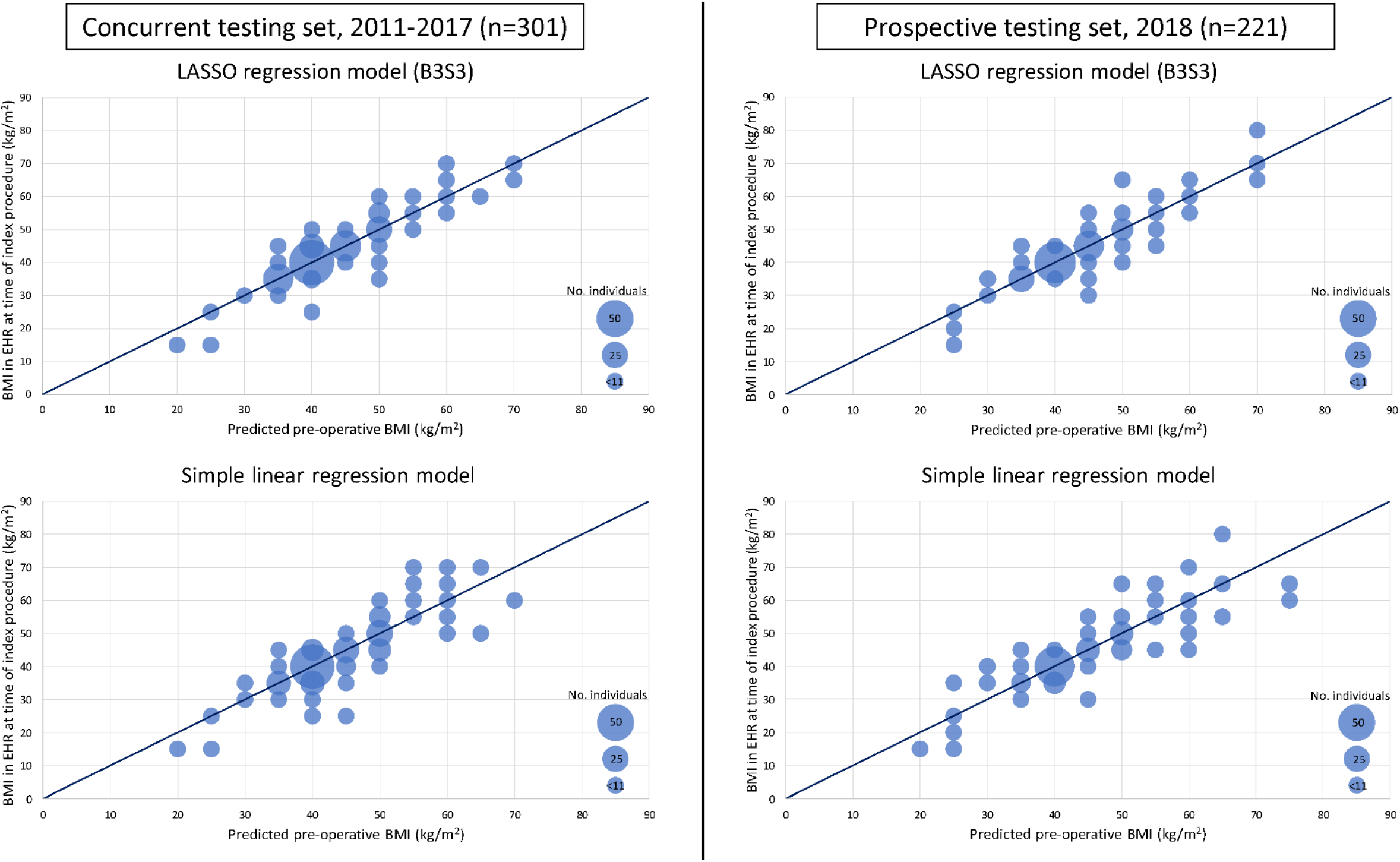
Calibration plots for the B3S3 and the simple linear regression model. Abbreviations: B3S3 = BMI Before Bariatric Surgery Scoring System; BMI = body mass index; EHR = electronic health record; LASSO = least absolute shrinkage and selection operator The calibration plots show the number of individuals falling within each 5-unit category of observed versus predicted BMI, where the size of the dots is proportional to the number of individuals in the category. Areas of the plot with <11 patients are represented using the same sized dots to maintain the de-identification nature of the database. Perfect calibration occurs along the diagonal line, where the predicted and observed BMI values are equivalent.

**Table 4.**
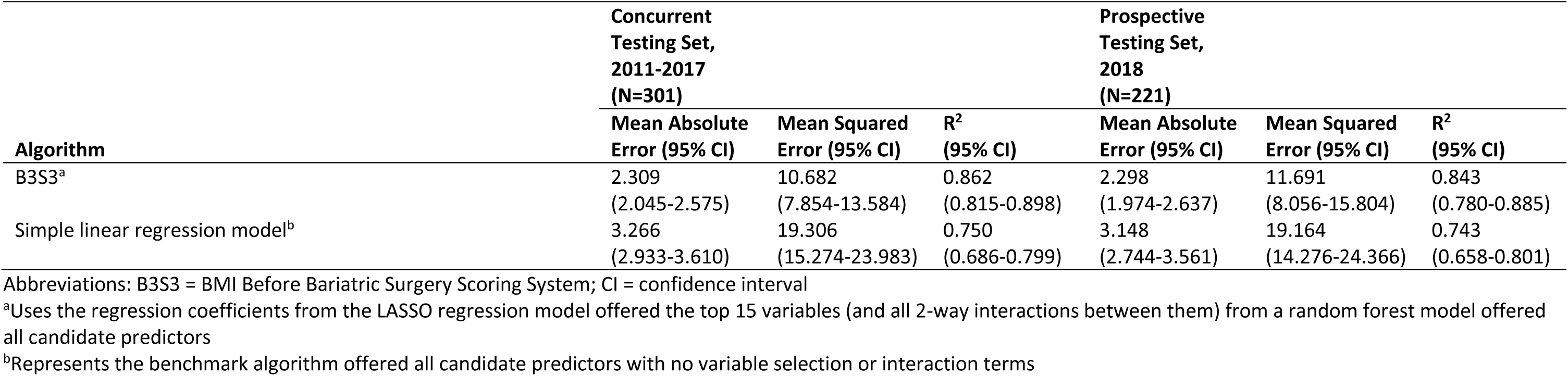
Calibration of the B3S3 and the simple linear regression model.

## DISCUSSION

To our knowledge, this study is the first to create a claims-based model to predict pre-operative BMI among bariatric surgery patients. By applying flexible, machine-driven model specification techniques within a carefully crafted machine learning pipeline, we derived a model that could accurately predict pre-operative BMI, as documented in the EHR, and be easily implemented as a simple scoring system with claims data without requiring any knowledge or experience with more complex machine learning algorithms, thus representing an accessible tool to all bariatric surgery researchers using claims data. This scoring system, the B3S3, which we have made publicly available along with instructions and an example of how to implement it (see Supplementary File 3), uses only a handful of claims-based variables, and its predictions were found to be highly accurate, even when evaluated in prospective data. Moreover, the B3S3 outperformed a linear regression model that utilized considerably more claims-based predictors but was not specified using a flexible, machine-guided process.

An important potential baseline confounder of post-surgical outcomes, the ability to accurately measure pre-operative BMI is an important consideration when identifying suitable databases for comparative effectiveness and safety research on bariatric surgeries. Many studies to date have used data from EHRs and bariatric surgery registries^5,7–10^ for their capture of more granular, clinical data, but claims data also represent a valuable data source for their more complete capture of numerous effectiveness and safety outcomes for bariatric procedures (e.g., cardiovascular events, abdominal operations, venous thromboembolism, and all-cause hospitalizations), particularly over longer follow-up horizons.^6^ However, as claims databases lack clinical measurements on BMI, our scoring system may enhance and promote the utility of claims data for bariatric surgery research in several ways. First, compared to the approach of imputing pre-operative BMI solely based on the descriptions of weight-related diagnosis codes (which only denote non-specific weight categories, or at best, *ranges* of BMI), the B3S3 allows researchers to predict pre-operative BMI as a *continuous* value and with the same level of granularity as BMI measurements typically recorded in EHRs and bariatric surgery registries. Thus, our scoring system allows researchers to enjoy maximum flexibility in how pre-operative BMI is handled in claims-only studies (e.g., as a categorical *or continuous* covariate). Second, as the B3S3 was found to be well calibrated along the full range of its predictions, these findings suggest that the B3S3 may be used to accurately identify cohorts of higher-risk bariatric surgery patients with extremely high pre-operative BMIs (e.g., ≥50 or ≥60 kg/m^2^) or exclude patients with lower than expected pre-operative BMIs (e.g., ≤35 kg/m^2^) whose operation may not be a true primary bariatric operation (e.g., revisional surgery) but otherwise appears as such based on other claims-based inclusion criteria. Third, even in EHR-based studies, the B3S3 may be useful as a remedy for missing data on pre-operative BMI when linked claims data are available. Indeed, a previous study^5^ had to exclude 16% of bariatric surgery patients due to lack of baseline data on BMI in the EHR. Finally, as pre-operative BMI has been found to be a significant predictor of post-operative outcomes,^19,23,35^ the B3S3 may also be useful in claims-based studies aimed at forecasting outcomes after bariatric surgery by enabling the creation of pre-operative BMI as a candidate prognostic variable.

The capacity of the B3S3 to accurately predict pre-operative BMI is largely due to 2 factors: 1) the existence of weight-related codes in the ICD system, many of which denote BMI-specific ranges, and 2) reimbursement requirements by health insurers that strongly incentivize providers and institutions to document weight-related codes for nearly all bariatric patients in the months preceding their index surgery. However, it is important to note that the validity of the B3S3 is most likely specific to bariatric surgery patients in the pre-operative period and may not be generalizable to the post-operative period (e.g., to measure post-operative weight loss), when providers and institutions no longer have financial incentives to document weight-related codes. Indeed, a prior study^16^ found that in the first post-operative year, one-fifth of bariatric surgery patients did not have any weight-related codes in their claims data and less than one-half of patients had BMI-specific codes documented. Thus, in bariatric surgery research, the main utility of weight-related codes (and by extension, the B3S3) is to enable confounding control and investigation of effect modification by baseline severity of obesity for post-surgical outcomes that are well captured in claims data.

A strength of this study is the inclusion of a nationally representative sample of bariatric surgery patients with commercial or Medicare Advantage plans, who currently represent the majority of US individuals who undergo bariatric surgery.^36^ By extension, a limitation of this study is that we could not include bariatric patients with other types of health insurance plans (e.g., Medicaid) due to the nature of data available in the OLDW. Another limitation is the smaller size of the testing sets that were used to evaluate the B3S3. To address sampling variability in our testing sets, we bootstrapped the testing data to estimate uncertainty around all performance measures. To further evaluate the generalizability of the B3S3, we strongly encourage other researchers to validate the performance of our scoring system in other claims databases, including data sources that capture other (e.g., Medicaid) patient populations.

## CONCLUSION

Pre-operative BMI is an important potential confounder in comparative effectiveness studies of bariatric surgeries. Although claims databases lack clinical measurements, the existence of weight-related diagnosis codes, combined with BMI-related reimbursement requirements by health insurers for bariatric surgeries, create valuable information on pre-operative BMI in claims data. We applied a machine learning pipeline to create a scoring system, the B3S3, that could accurately predict pre-operative BMI, as documented in the EHR, among bariatric surgery patients. Investigators can easily use the B3S3 with claims data to obtain granular predicted values of pre-operative BMI to enhance confounding control and investigation of effect modification by baseline severity of obesity in bariatric surgery studies.

## Data Availability

The raw data that support the findings of this study are available from Optum Labs, but restrictions apply to the availability of these data, which were used under license for the current study and so are not publicly available.

## LIST OF ABBREVIATIONS

AGB: Adjusted gastric banding
BMI: Body mass index
B3S3: BMI Before Bariatric Surgery Scoring System
CI: Confidence interval
EHR: Electronic health record
ICD: International Classification of Diseases
LASSO: Least absolute shrinkage and selection operator
MAE: Mean absolute error
MSE: Mean squared error
OLDW: Optum Labs Data Warehouse
RYGB: Roux-en-Y gastric bypass
SG: Sleeve gastrectomy

## DECLARATIONS

### Ethics approval and consent to participate

This study was reviewed and approved by the Harvard Pilgrim Health Care Institutional Review Board with a waiver of individual patient consent. All methods in this study were carried out in accordance with the relevant ethical guidelines and local regulations.

### Consent for publication

Not applicable.

### Availability of data and materials

The raw data that support the findings of this study are available from Optum Labs, but restrictions apply to the availability of these data, which were used under license for the current study and so are not publicly available. The materials generated for the current study are included in this published article and its supplementary files.

### Authors’ contributions

JW conceived and designed the work, performed the analysis, interpreted the data, and drafted the manuscript; XL created the datasets used in the work, interpreted the data, and reviewed/edited the manuscript; DEA contributed to the design of the work, interpreted the data, and reviewed/edited the manuscript; DL interpreted the data and reviewed/edited the manuscript; EMJ interpreted the data and reviewed/edited the manuscript; RW contributed to the design of the work, interpreted the data, and reviewed/edited the manuscript; and ST acquired the data, contributed to the conception and design of the work, interpreted the data, and reviewed/edited the manuscript. All authors have read and approved the final manuscript and agree to be accountable for their contributions, the accuracy, and the integrity of the work.

## Acknowledgements

The authors would like to thank Dr. Susan Gruber for her helpful input on the analytic approach used in this study.

