## Supplementary File 1 for "Using claims data to predict pre-operative BMI among bariatric surgery patients: development of the BMI Before Bariatric Surgery Scoring System (B3S3)"

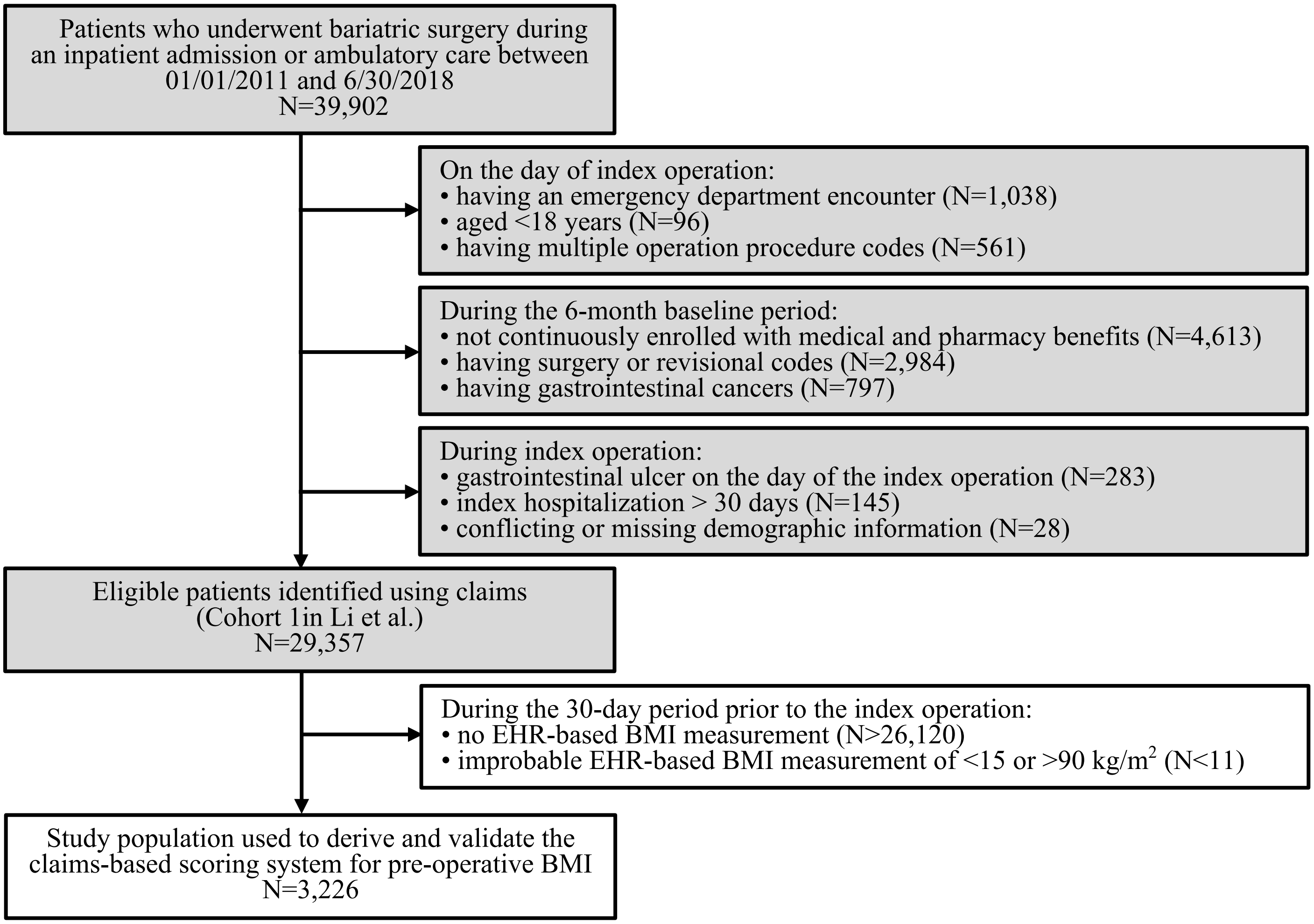


**eFigure 1. Inclusion and exclusion criteria used to identify the study population**

We replicated the inclusion and exclusion criteria for *Cohort 1* in Li *et al.*,[1] which was used to evaluate the availability of weight-related diagnosis codes during the preoperative and postoperative periods in a retrospective cohort of bariatric surgery patients (grey colored boxes). From among the eligible patients in *Cohort 1*, we identified the subset of bariatric surgery patients with a BMI measurement recorded in their linked EHR data on or within 30 days prior to the day of the index procedure, where BMI values <15 or >90 kg/$m^2$ were excluded to avoid including probable data entry errors (white colored boxes). This subset of bariatric surgery patients comprised the final study population that we used to derive and validate the claims-based scoring system for pre-operative BMI. Note that exclusion criteria with <11 individuals have been masked to maintain the deidentification nature of the database.
