## Supplementary File 2 for "Using claims data to predict pre-operative BMI among bariatric surgery patients: development of the BMI Before Bariatric Surgery Scoring System (B3S3)"

### eTable 1. Code mappings for weight-related diagnosis codes into weight categories

| Weight Category[a] | ICD-9-CM Codes | ICD-10-CM Codes |
|---|---|---|
| Underweight, normal weight, or overweight | 78322,<br>V850,<br>V851,<br>V8521, V8522, V8523, V8524, V8525,<br>27802 | R636,<br>Z681<br>Z6820, Z6821, Z6822, Z6823, Z6824,<br>Z6825, Z6826, Z6827, Z6828, Z6829,<br>E663 |
| BMI 30.0-34.9 | V8530, V8531, V8532, V8533, V8534 | Z6830, Z6831, Z6832, Z6833, Z6834 |
| BMI 35.0-35.9 | V8535 | Z6835 |
| BMI 36.0-36.9 | V8536 | Z6836 |
| BMI 37.0-37.9 | V8537 | Z6837 |
| BMI 38.0-38.9 | V8538 | Z6838 |
| BMI 39.0-39.9 | V8539 | Z6839 |
| BMI 40.0-44.9 | V8541 | Z6841 |
| BMI 45.0-49.9 | V8542 | Z6842 |
| BMI 50.0-59.9 | V8543 | Z6843 |
| BMI 60.0-69.9 | V8544 | Z6844 |
| BMI ≥70.0 | V8545 | Z6845 |
| Obese, nonspecific | 27800,<br>27803 | E669,<br>E6609,<br>E661,<br>E668 |
| Severely obese, nonspecific | 27801 | E6601,<br>E662 |

Abbreviations: BMI = body mass index; ICD-9-CM = International Classification of Diseases, Ninth Revision, Clinical Modification; ICD-10-CM = International Classification of Diseases, Tenth Revision, Clinical Modification

[a]Each weight category was a unique candidate predictor in the claims-based models and scoring system, where weight categories were not mutually exclusive (to capture patients who had ICD codes for multiple weight categories documented in the pre-operative period).

**eTable 2. Tuning parameters for the random forest and LASSO regression models**

| Algorithm | Tuning Parameter | Description | Subset of values considered[a] | Tuned value[b] |
|---|---|---|---|---|
| Random forest | nTree | Number of trees | {50, 100, 250, 500, 1000, 1500, 2000} | 1000 |
| | mTry | Number of candidate features considered at each node | {5, 10, 15, 30, 60} | 15 |
| LASSO | lambda | Controls amount of regularization (shrinkage) applied to the model coefficients | Sequence of 100 values automatically selected by the glmnet package in R | 0.0283 |

Abbreviations: LASSO = least absolute shrinkage and selection operator

[a]A 10-fold cross-validation procedure in the training set was used to evaluate the (cross-validated) performance of each algorithm across the subset of values for the tuning parameter. For the random forest, a grid of all possible combinations for the tuning parameter values were evaluated (7 values for nTree X 5 values for mTry = 35 combinations).

[b]The tuned values were used in the final fitted models and represent the value (or set of values for the random forest) that yielded the algorithm with the lowest cross-validated mean squared error in the training set.

### eTable 3. Full set of comorbidities considered for prediction models

| Comorbidity n (%) | Overall (N=3,226) | Training Set, 2011-2017 (N=2,704)[a] | Concurrent Testing Set, 2011-2017 (N=301)[a] | Prospective Testing Set, 2018 (N=221)[a] |
|---|---|---|---|---|
| Hypertension | 2236 (69.3) | 1871 (69.2) | 219 (72.8) | 146 (66.1) |
| Gastroesophageal reflux disease | 1942 (60.2) | 1614 (59.7) | 191 (63.5) | 137 (62.0) |
| Sleep apnea | 1826 (56.6) | 1516 (56.1) | 181 (60.1) | 129 (58.4) |
| Dyslipidemia | 1825 (56.6) | 1534 (56.7) | 169 (56.2) | 122 (55.2) |
| Diabetes | 1159 (35.9) | 952 (35.2) | 118 (39.2) | 89 (40.3) |
| Depression | 1115 (34.6) | 926 (34.3) | 112 (37.2) | 77 (34.8) |
| Anxiety | 1080 (33.5) | 905 (33.5) | 95 (31.6) | 80 (36.2) |
| Liver disease | 875 (27.1) | 718 (26.6) | 93 (30.9) | 64 (29.0) |
| Chronic pulmonary disease | 841 (26.1) | 675 (25.0) | 97 (32.2) | 69 (31.2) |
| Psychosis | 830 (25.7) | 680 (25.2) | 67 (22.3) | 83 (37.6) |
| Non-alcoholic fatty liver disease | 825 (25.6) | 681 (25.2) | 87 (28.9) | 57 (25.8) |
| Acquired hypothyroidism | 635 (19.7) | 519 (19.2) | 68 (22.6) | 48 (21.7) |
| Complicated diabetes | 494 (15.3) | 389 (14.4) | 43 (14.3) | 62 (28.1) |
| Deficiency anemia | 472 (14.6) | 400 (14.8) | 40 (13.3) | 32 (14.5) |
| Cardiac arrhythmias | 464 (14.4) | 383 (14.2) | 40 (13.3) | 41 (18.6) |
| Eating disorder | 373 (11.6) | 316 (11.7) | 36 (12.0) | 21 (9.5) |
| Congestive heart failure | 302 (9.4) | 250 (9.3) | 24 (8.0) | 28 (12.7) |
| Osteoarthritis, lower limb | 298 (9.2) | 242 (9.0) | 26 (8.6) | 30 (13.6) |
| Fluid and electrolyte disorders | 288 (8.9) | 239 (8.8) | 26 (8.6) | 23 (10.4) |
| Kidney diseases | 242 (7.5) | 203 (7.5) | 17 (5.7) | 22 (10.0) |
| Substance use disorder | 185 (5.7) | >153 (>5.7) | <11 (<3.7) | 21 (9.5) |
| Psychotic disorder | 182 (5.6) | 158 (>5.8) | 13 (4.3) | <11 (<5.0) |
| Renal failure | 171 (5.3) | >143 (>5.3) | <11 (<3.7) | 17 (7.7) |
| Peripheral vascular disease | 160 (5.0) | 136 (5.0) | 11 (3.7) | 13 (5.9) |
| Polycystic ovarian syndrome | 148 (4.6) | >126 (>4.7) | <11 (<3.7) | <11 (<5.0) |
| Smoker | 134 (4.2) | 119 (4.4) | 15 (5.0) | 0 (0.0) |
| Any tumor | 131 (4.1) | >104 (>3.8) | <11 (<3.7) | 16 (7.2) |
| Weight loss | 110 (3.4) | >88 (>3.3) | 11 (3.7) | <11 (<5.0) |
| Pulmonary circulation disorders | 89 (2.8) | >64 (>2.4) | <11 (<3.7) | 14 (6.3) |
| Coagulopathy | 78 (2.4) | >56 (>2.1) | <11 (<3.7) | <11 (<5.0) |
| Deep vein thrombosis | 55 (1.7) | >33 (>1.2) | <11 (<3.7) | <11 (<5.0) |
| Pulmonary embolism | 46 (1.4) | >24 (>0.9) | <11 (<3.7) | <11 (<5.0) |
| Alcohol abuse | 35 (1.1) | >24 (>0.9) | <11 (<3.7) | 0 (0.0) |

[a]Cells with <11 patients have been masked to maintain the deidentification nature of the database.
